# SENSOR TECHNOLOGY FOR OPENING NEW PATHWAYS IN DIAGNOSIS AND THERAPEUTICS OF BREAST, LUNG, COLORECTAL AND PROSTATE CANCER

**DOI:** 10.1101/2022.02.18.22271186

**Authors:** Saeed Roshani, Mario Coccia, Melika Mosleh

## Abstract

This study analyzes the interaction between sensor research and technology and different types of cancer (breast, lung, colorectal and prostate) that generates a high mortality in society with the goals of detecting new directions for improving diagnosis and therapeutics. This study develops an approach of computational scientometrics based on data of the Web of Science from 1991 to 2021 period. The results of this analysis show the vital role of biosensor and electrochemical biosensor applied in breast cancer, lung cancer, and prostate cancer research, except colorectal cancer for latter one. Instead, research of optical sensors is developing mainly in the directions of breast, prostate, and colorectal cancer for improving diagnostics. Finally, oxygen sensor research is developing for breast and lung cancer for applications in breath analysis in new treatment processes. Preliminary results presented here clearly illustrate the evolutionary paths of main sensor technologies that have a great potential in future diagnosis of cancer, but we also need additional examinations of other aspects and factors for supporting appropriate strategies of management of technology to foster the technology transfer of sensor technologies in cancer research for improving diagnosis and, whenever possible, reduce world-wide mortality of cancer ins society.

## 1. Introduction and goal of investigation

The research field of sensor is undergoing a significant change to support the evolution of science and technologies in society (Andersen et al., 2004; Coccia et al., 2021; Coccia and Watts, 2020; Coccia, 2019, 2019a, 2021, 2021a; Rao et al., 2018; Wilson, 2004). The goal of this study is an exploratory analysis to detect main sensor technologies applied in cancer research for improving diagnosis and treatments and reducing whenever possible mortality between countries. The vast literature in these topics shows main results for cancer research (Bayford et al., 2022; Kaur et al., 2022; Li et al., 2020, 2020a; Rey-Barth et al., 2022; Sivanandhan et al., 2022; Thakare et al., 2022). As far as breast cancer is concerned, Wu et al. (2022) design a dual-aptamers functionalized gold for classification of breast cancer based on Förster resonance energy transfer, which is potentially useful for quantitative classification of different subtypes of breast cancer. Lu et al. (2022) argue that phthalates can penetrate the environment and enrich various aquatic organisms through the food chain, which is involved in promoting the growth of breast cancer. It is of current interest to develop new sensors for phthalates. Results show that guest-induced reassembly brings forth significant fluorescence change, which is a promising way of designing new fluorescent probes for the analysis of phthalates in the environment and food. Pothipor et al. (2022) show that a dual-mode electrochemical biosensor is successfully developed for simultaneous detection of two different kinds of breast cancer biomarkers. The experimental results suggest that this label-free biosensor exhibits good linear responses to the concentrations of both target analytes with the limits of detection. This assay strategy has a great potential to be further developed for the simultaneous detection of a variety of miRNAs and protein biomarkers for point-of-care diagnostic applications. Kim et al. (2022) maintain that mechanophores are molecular motifs that respond to mechanical perturbance with targeted chemical reactions toward desirable changes in material properties. Taking advantage of the strengths of mechanophores and high-intensity focused ultrasound, mechanochemical dynamic therapy can provide noninvasive treatments for diverse cancer types (cf., Mohan et al., 2022). About prostate cancer, Bax et al. (2022) argue that diagnostic protocol is affected by poor accuracy and high false-positive rate and propose an electronic nose for non-invasive prostate cancer detection. The approach proved to be effective in mitigating drift on 1-year-old sensors by restoring accuracy from 55% to 80%, achieved by new sensors not subjected to drift. The model achieved, on double-blind validation, a balanced accuracy of 76.2%. Prema et al. (2022) examine the biological synthesis of gold nanoparticles using green tea and their cytotoxicity against human prostate cancer cells. The findings suggest that the biosynthesized reduced prostate cancer cell proliferation and exert their anti-proliferative action on the prostate cancer cell line by inhibiting growth, decreasing DNA synthesis, and triggering apoptosis. In lung cancer, Joshi et al. (2022) argue that proper and early diagnosis of cancers are a basic to efficient treatment and better prognosis and report a simple and label-free method of detection of two antigens: carcinoembryonic antigen (CEA) and cytokeratin-19 fragment (CYFRA 21-1) that are the biomarkers of many cancers including lung cancer. The responses of the sensors ranged from 10.96 to 26.48% for 0.25 pg/mL to 20 ng/mL CEA and it varied from 17.66 to 26.68% for 0.25 pg/mL to 20 ng/mL CYFRA 21-1. Kaya et al. (2022) review the recent advances and improvements (2011–2021 period) in nanomaterials based electrochemical biosensors for the detection of the lung and colon cancer biomarkers (cf., Tumuluru et al., 2022). In colon cancer, Jiang et al. (2022) argue that Transient receptor potential vanilloid 1 (TRPV1) acts as cellular sensor and is implicated in the tumor microenvironment cross talk and the functional role of TRPV1 in colorectal cancer (CRC). The study reveals an important role for TRPV1 in regulating the immune microenvironment during colorectal tumorigenesis and might be a potential target for CRC immunotherapy. Welz et al. (2022) point out that the intestinal epithelium undergoes constant self-renewal from intestinal stem cells. Together with genotoxic stressors and failing DNA repair, this self-renewal causes susceptibility toward malignant transformation. The study shows that X-box binding protein 1 is a stress sensor involved in coordinating epithelial DNA damage responses and stem cell function.

In this context of the evolution of sensor technologies towards manifold fields of research, the motivation of this study is to clarify the role of research and technology in sensors for main typologies of cancer, describing the networks of interconnection of sensors with other technologies and scientific aspects related to cancer under study. Proposed methodology can indicate new directions of sensor technologies for diagnosis and treatments of cancer and help policymakers to allocate with efficiency financial resources to support scientific and technological development in these critical fields of research in society (cf., also Ardito et al., 2021; Coccia, 2018, 2019a, 2020; Coccia and Finardi, 2012, 2013; Kashani and Roshani, 2019; Roshani et al., 2021).

The balance of the paper proceeds as follows. First, it describes the data and methodology, applying a novel information processing approach of computational scientometrics, to generate maps of science that can explain the ecosystem and evolution of sensor research and technologies in specific typologies of cancer. We then show the results and conclude with a discussion on new directions of the evolution of sensor technology in cancer and limitations of the study to be solved with future studies.

## 2. Study design

First, this study focuses on main typologies of cancer that have the highest estimated age-standardized incidence and mortality rate worldwide as indicated in Table 1 and Figure 1 based on data by Globocan (2020).

**Table 1.** Estimated age-standardized incidence and mortality rates (World) in 2020, worldwide, both sexes, all ages

| Cancer | Incidence | Mortality |
| --- | --- | --- |
| Breast | 47.8 | 13.6 |
| Prostate | 30.7 | 7.7 |
| Lung | 22.4 | 18 |
| Colorectum | 19.5 | 9 |
| Cervix uteri | 13.3 | 7.3 |
| Stomach | 11.1 | 7.7 |
| Liver | 9.5 | 8.7 |
| Corpus uteri | 8.7 | 1.8 |
| Ovary | 6.6 | 4.2 |
| Thyroid | 6.6 | 0.43 |

Data source: Globocan (2020)

**Figure 1.**
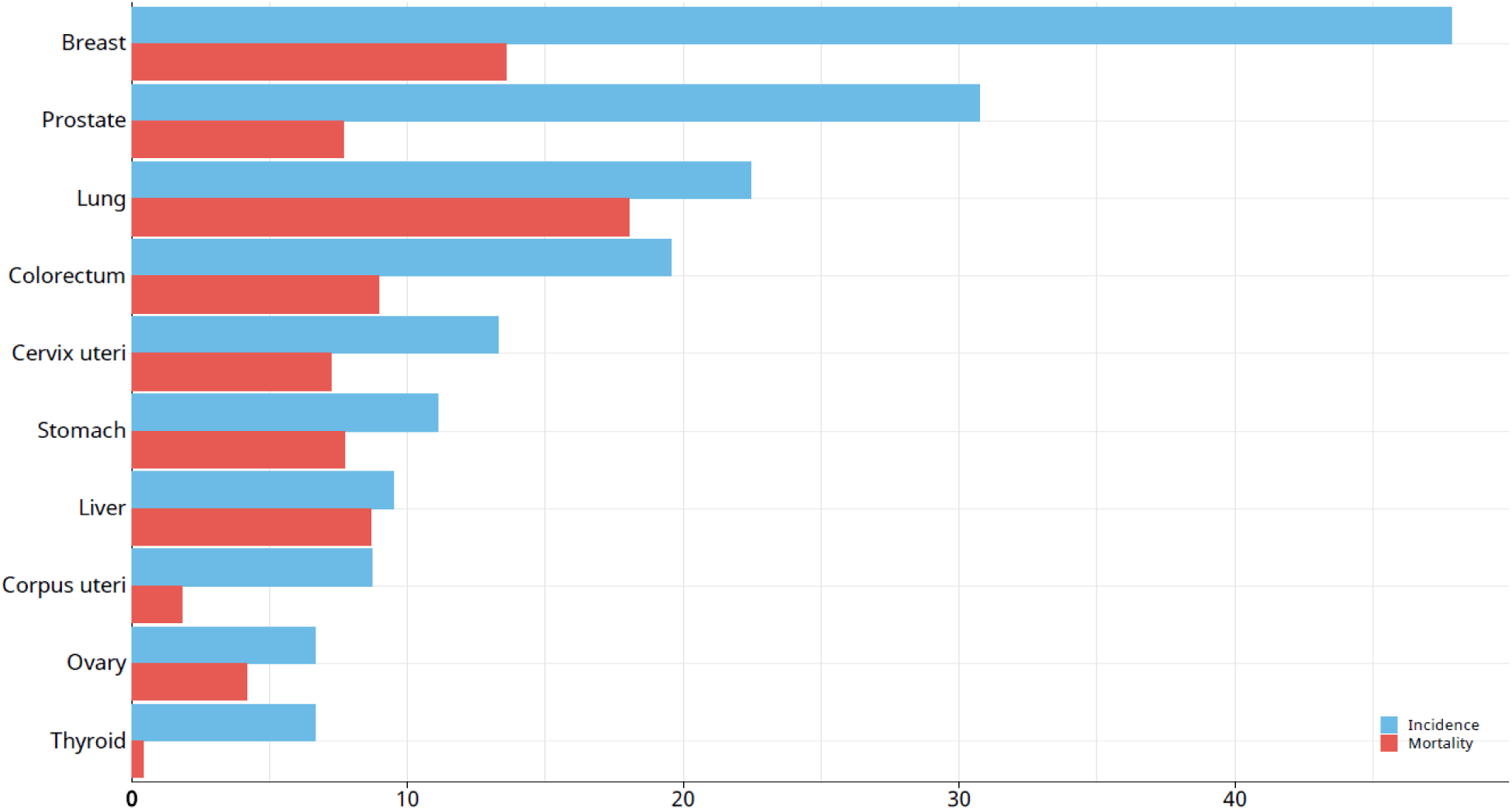
Estimated age-standardized incidence and mortality rates (World) in 2020, worldwide, both sexes, all ages. Red bars indicate mortality, blue bares are incidence. Data source: Globocan (2020)

Considering the data just mentioned, this study focuses on the investigation of sensor technology and research in the following four main types of cancer:

- Breast cancer
- Lung cancer
- Prostate cancer
- Colorectal cancer

### 2.1 Data sources and retrieval strategy

To address the main question of this study, we used the Web of Science (WOS, 2021) core collection database to retrieve the articles related to sensor technologies and cancers. In this study, we focused as said on four major types of cancers for a comparative analysis (Coccia, 2018a) including: Breast cancer (BC), lung cancer (LC), Prostate cancer (PC) and Colorectal cancer (CC). We used the following strategy for extracting related articles for further processing. The term “sensor” was searched with “breast cancer” in the topics of articles. We changed the “breast” to “lung” for extracting the articles related to lung cancer, “prostate” for prostate cancer and “colorectal” for colorectal cancer. The results are refined by document type = “Articles”, Language = “English”, Publication years = (1991-2021), Web of Science index = “SCI-EXPANDED”). We found 1117 unique articles for breast cancer, 764 articles for lung cancer, 454 articles for prostate cancer, and 282 articles for colorectal cancer.

### 2.2 Data processing procedure

To find the applications of sensors technology in cancers considered in this article, we used the original keywords (DEs) provided by the authors as the basis for constructing the word co-occurrences networks. According to this technique, two terms are considered co-occurrence whenever they simultaneously appear in a single document (Delecroix and Epstein, 2004). To find the relevant sensor technologies to each of the cancer we studied, we applied the following procedure:

#### Keywords standardization

We tried to clean the keywords according to their meaning and structures in this step. For instance, we combined the “bio-sensor” and “biosensor” into Biosensor. Also, we changed all abbreviations and plural forms of nouns into a basic form (e.g., computers changed to computer and “AMPK” changed to “Amp-activated Protein kinase”.

#### Network construction

We used SCI2 tool V. 1.3 for constructing the word co-occurrences network (Sci2 Team, 2009). As mentioned above, we used the article’s original keywords (DE tag) to create the networks. We create four different breast, lung, Prostate, and colorectal cancer networks. Also, we removed the isolated nodes after analyzing the nodes and links in the networks.

#### Path-finding

path-finding is a technique for choosing the shortest links between two nodes. To reduce the links of our networks and emphasize the most important nodes, we used a minimum spanning tree (MST) path-finder algorithm (Quirin et al., 2008). All the calculations are implemented by SCI2 tool. Also, as an input parameter of the algorithm, we set the parameter Weight Attribute measures to “SIMILARITY” and Edge Weight Attribute to “Unweighted”. The initial network of breast cancer contains 10,149 links, and links reduced network contains 2,318 edges. The lung cancer network contains 6,128 initially and has 1539 links after applying the link reduction algorithm. After the algorithm implementation, the Prostate cancer network had 3,395 links and held 850 edges. These results for colorectal cancer include 2,394 links at the first and 528 links after implementing the link reduction algorithm.

#### Visualizations

We utilized Gephi software (Bastian et al., 2009) version 0.9.2 to visualize the networks The nodes indicate the original keywords, and links show the co-occurrences among them. Also, the size of nodes is based on the Betweenness centrality. This measure is used for detecting the nodes which have a connecting role in the network (Kashani and Roshani, 2019). We set the Betweenness centrality as an indicator for identifying the groups of the path in each network. In other words, nodes with Betweenness centrality greater than 0.1 are considered thresholds for identifying the groups.

## 3. Results

### 3.1. Breast cancer

Breast cancer has seriously threatened women health in the world (Chagpar and Coccia, 2019; Coccia, 2013, 2019c,1029b, 2020; Harbeck and Gnant, 2017). As mentioned earlier, the breast cancer sample contains 1,117 articles, and it is the most extensive dataset in our analysis. This network contains 2,319 nodes (keywords) and 2,318 links. Also, this network includes 149 sensors that are interconnected to the other nodes. Figure 2 shows the breast cancer co-word network.

**Figure 2.**
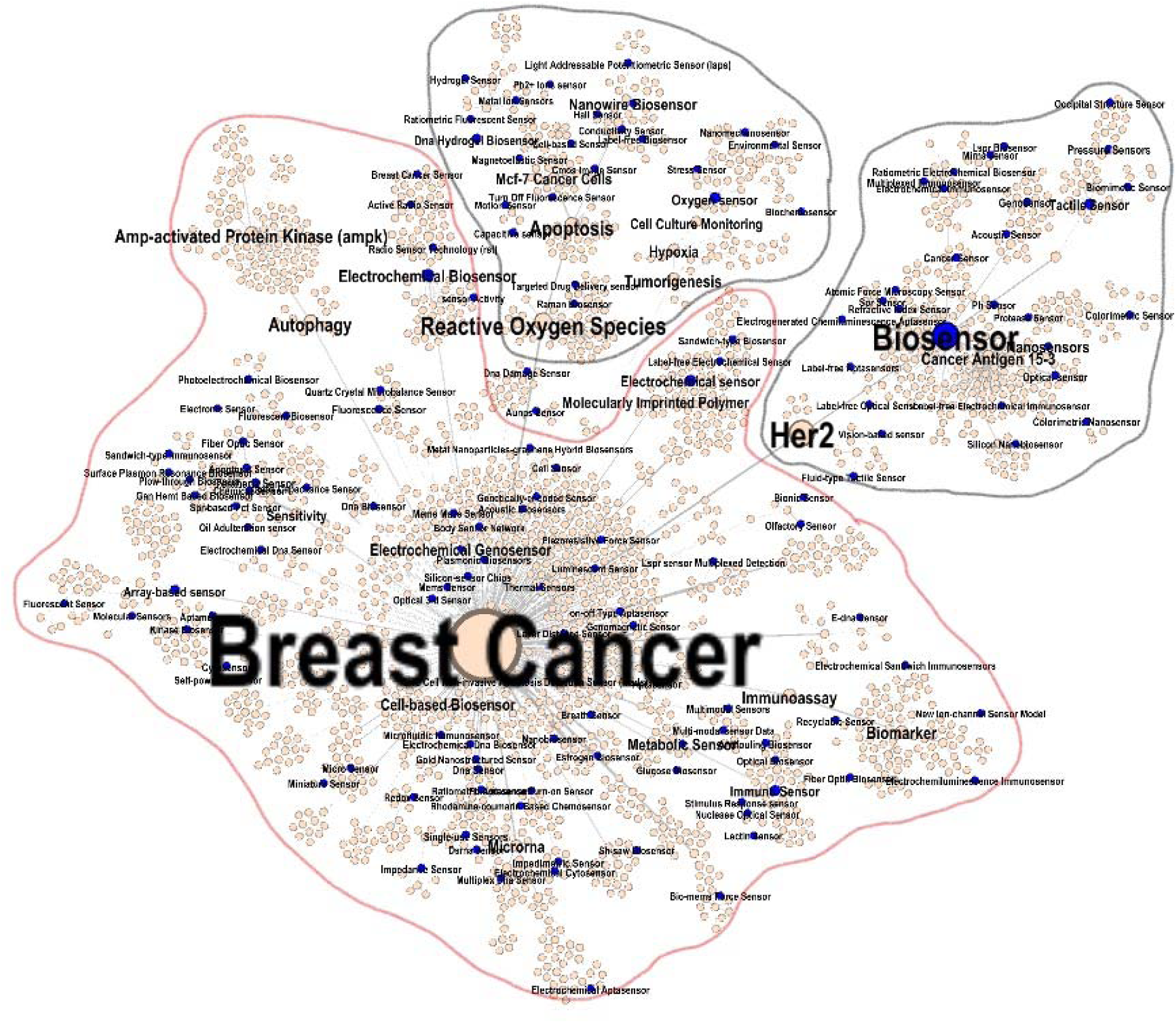
Co-word analysis map of breast cancer.

Based on the Betweenness centrality value, we found three sub-groups in this network. Table 1A in Appendix shows the most important information about these groups. Also, there are 149 sensors interconnected to this network.

In group 1 related to breast cancer, we have five hot topics: Autophagy, Immunoassay, Electrochemical Biosensor, Tumorigenesis, and Microrna, which have a high co-occurrence with Electrochemical Biosensor, Electrochemical Sensor, Oxygen Sensor, Immuno sensor, and Array-based Sensor.

Group 2 with head of Biosensor has a couple of hot topics, including Her2, Cancer antigen, Nanoparticle, Tactile Sensor, and Signal Amplification, which are significantly related to Optical sensor, Colorimetric Sensor, Colorimetric Nanosensor, Ph. Sensor, and Refractive Index Sensor.

Group 3 led by Reactive Oxygen Species, it is related to Apoptosis, Mcf-7 Cancer Cells, Circulating Tumor Cell, Dna Hydrogel Biosensor, and Carbon Dot. In this domain, Raman Biosensor, DNA Hydrogel Biosensor, Light Addressable Potentiometric Sensor (laps), Capacitive Sensor, and Label-free Biosensor are couple of sensors which is included in this group of connection (see in Appendix Table 1A about groups, core keywords and related sensors in the breast cancer network).

### 3.2. Lung cancer

According to the size of the network, the lung cancer is the second cancer were analyzed (cf., Coccia, 2012; 2014; 2017; 2019d). This network contains 1,540 nodes (keywords) and 1,539 links. Also, this network includes 121 sensors that interconnected to the other nodes. Figure 3 shows the lung cancer co-word network.

**Figure 3.**
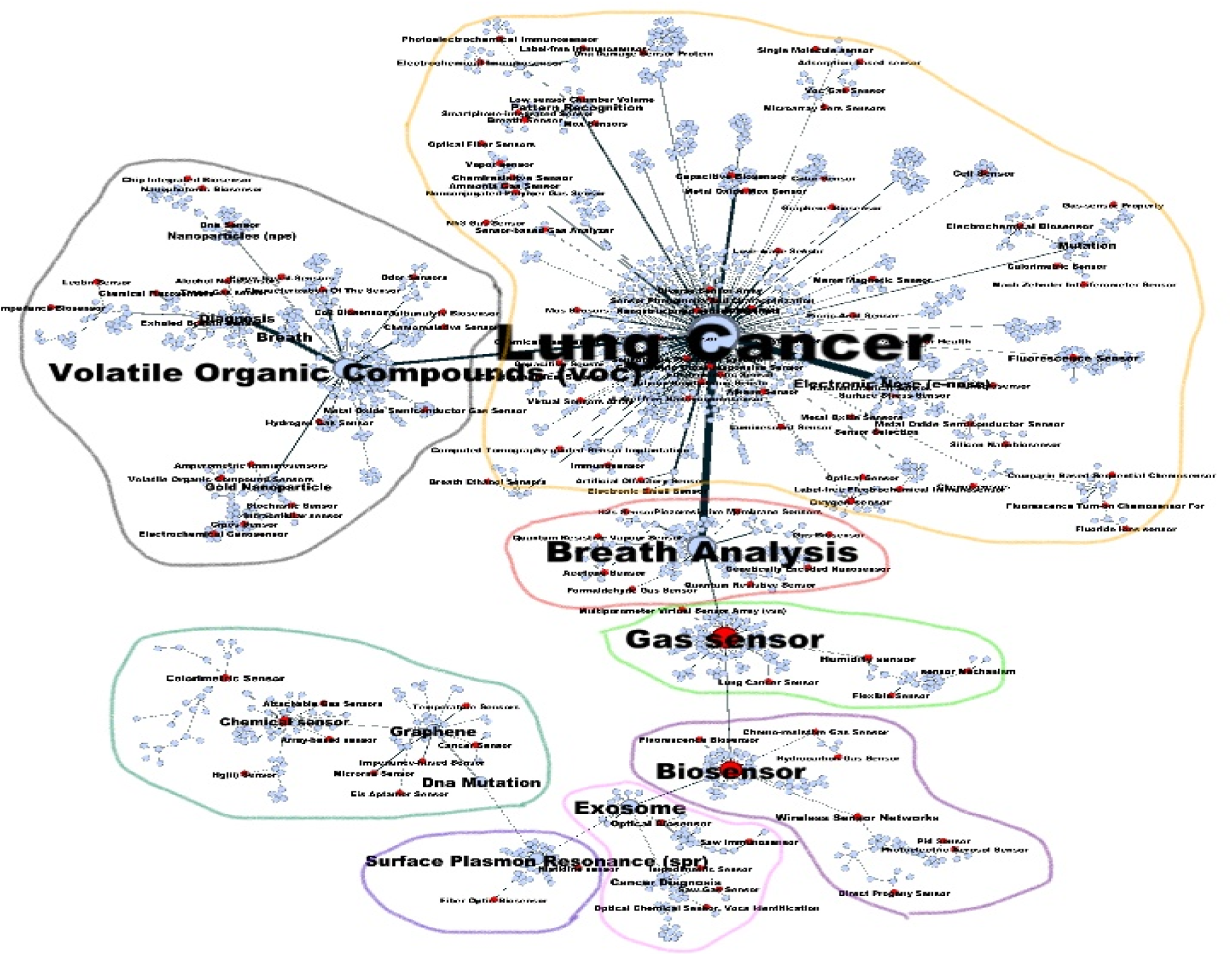
Co-word analysis map of lung cancer

As shown in figure 3, Regarding the Betweenness centrality threshold of 0.1, this network has eight path groups. All nodes with a value of Betweenness centrality greater than 0.1 are considered the head of a group. Top keywords and all the sensors included in these categories have been shown in Table 2A in Appendix.

Group 1 with the head of lung cancer is related to 5 hot topics: Electronic Nose (e-nose), Pattern Recognition, Mutation, Nrf2 (Nuclear factor-erythroid factor 2-related factor 2), and Calix[4]arene. It is also illustrated that 69 sensors are interconnected to this path. The oxygen sensor, Electrochemical Biosensor, Cell Sensor, Metal Oxide Semiconductor Sensor, Chemiresisitve Sensor, Breath Sensor, Electrochemical Immunosensor, and Capacitive Biosensor are a couple of sensors that are involved in the creation of linkage path among these nodes.

In group 2 led by Volatile Organic Compounds (voc), we identified 23 related sensors including, Exhaled Breath Sensor, Dna Sensor, Volatile Organic Compound Sensors, Cmos Sensor, Electrochemical Genosensor, etc. The hot topics related to this group of nodes are Breath, Diagnosis, Gold Nanoparticle, Nanoparticles, and Exhaled Breath.

Group 3 connected to Breath analysis, has 8 interconnected sensors, including Piezoresistive Membrane Sensors, H2s Sensor, Formaldehyde Gas Sensor, Quantum Resistive Vapor Sens Quantum Resistive Sensor, Acetone Sensor, and Genetically Encoded Nanosensor. The most frequent keywords involved in this path group are: Real-Time, Diabetes, Non-invasive, Formaldehyde, and Sno2.

Group 4 with the head of Gas sensor is related to important keywords of Humidity sensor, Health Monitoring, Reduced Graphene Oxide, and Acetone. Six sensors of gas, lung cancer, Sensor, multiparameter virtual sensor array, humidity sensor, sensor Mechanism, and Flexible Sensor are part of this path group.

In Group 5, Biosensor with the most important technology in creating the path among different sensors and keywords, it is responsible for making a bridge between the other seven sensors of Fluorescence Biosensor, Wireless Sensor Networks, Photoelectric Aerosol Sensor, Direct Progeny Sensor, Pid Sensor, Chemo-resistive Gas Sensor and most frequent keywords of Wireless Sensor Networks, Radon, Smart Home, Electrochemical Inhibitors, and Hydrocarbon Gas Sensor.

The next group, led by Exosome connected to the top five keywords of Cancer Diagnosis, Optical Biosensor, Immunoassay, and Multi-wall Carbon Nanotubes related to five sensors of Optical Biosensor, Saw Gas Sensor, Optical Chemical Sensor, Vocs Identification, Saw Immunosensor, and Impedimetric Sensor.

The 7^th^ group distinguished by Surface Plasmon Resonance (spr) includes five hot keywords of Endoscopy, Au Nps, Signal Enhancement, Erlotinib, and Tollen’s Reagent, which have two sensors of Histidine sensor and Fiber Optic Biosensor in their category.

In group 8 related to Graphene, we have five DNA Mutation, Chemical Sensor, Urine Headspace, Colorimetric Sensor, and Tuberculosis keywords. Ten sensors of Chemical Sensor, Colorimetric Sensor, Cancer Sensor, Array-based Sensor, Temperature Sensors, Impedance-based Sensor, Attachable Gas Sensors, Hg(ii) Sensor, Eis Aptamer Sensor, and Microrna Sensor are the important sensing technologies in this group.

### 3.3. Prostate cancer

This network contains 870 nodes (keywords) and 869 links. This network includes 72 sensors that are interconnected to the other nodes. Figure 4 shows the prostate cancer co-word network.

**Figure 4.**
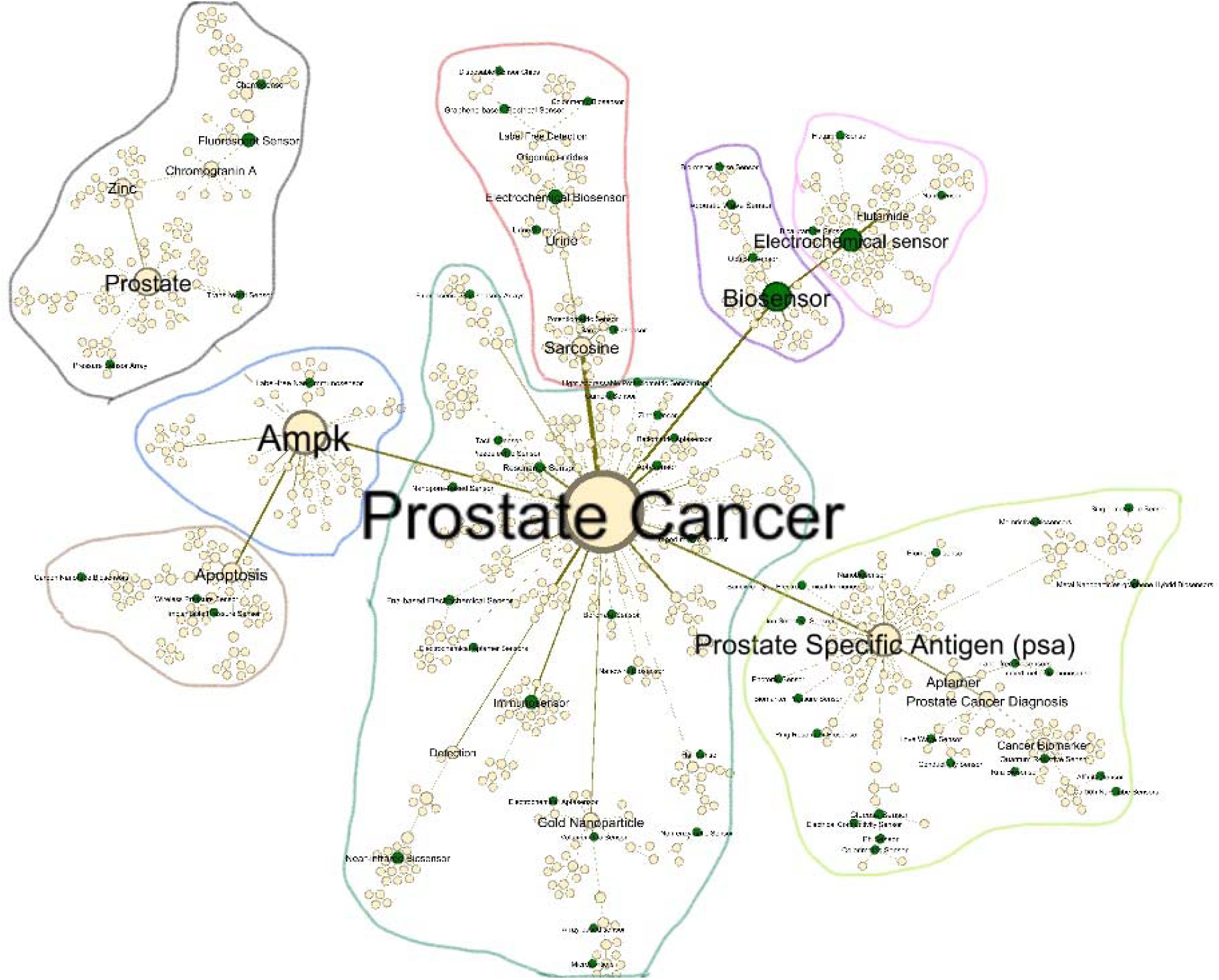
Co-word analysis map of Prostate cancer

Figure 4 shows that there are eight groups in the Prostate cancer network. The most important nodes based on the Betweenness centrality value are: Prostate cancer, Prostate-specific antigen, Activated Protein Kinase (AMPK) and Biosensor. Table 3A in Appendix shows the groups, core keywords of them and related sensors.

Group 1, led by Prostate cancer, is related to 5 hot topics of Gold Nanoparticle, Immunosensor, Detection, Porcine Liver Esterase, and Fluorescent Probe. It is also illustrated that 27 sensors are interconnected to this path. Immunosensor, Near-infrared Biosensor, Resonance Sensor, Fna-based Electrochemical Sensor, Impedimetric Sensor, and Aptasensor are a couple of sensors involved in the creation of linkage path among these nodes.

In group 2, led by Activated Protein Kinase (Ampk), we identified Label-free Nanoimmunosensor as the only connected Sensor. The hot topics related to this group of nodes are Apoptosis, Lkb1, Lung Cancer, Mtorc1, Castrate-resistant

Group 3, connected to Prostate Specific Antigen (PSA), has 22 interconnected sensors, including Glucose Sensor, Ph Sensor, Colorimetric Sensor, Quantum Resistive Sensor, etc. The most frequent keywords involved in this path group are Aptamer, Prostate Cancer Diagnosis, Cancer Biomarker, and Silicon Nanowire.

Group 4 with the head of Biosensor is related to important keywords of Antibody, Prostatic Carcinoma, DNA, Acoustic Wave Sensor, Cancer Metastasis. Four sensors of Biosensor, Optical Sensor, Acoustic Wave Sensor, Bio-mems Force Sensor are part of this group.

The next group, led by Prostate connected to the top five Zinc, Chromogranin A, Fluorescent Sensor, Imaging Diagnosis, and Peptide related to four sensors of Trans-rectal and Pressure Sensor Array, Chemosensor, and Fluorescent Sensor.

In Group 6, Electrochemical Sensor with the most important position in creating the path among different sensors and keywords is responsible for making a bridge between Nanosensor, Flutamide Sensor, Bicalutamide Sensor and most frequent keywords of Flutamide, Nanocomposite, Voltammetry, Anticancer Drug, and Electrophilicity.

The 7^th^ group distinguished by Sarcosine includes four hot keywords of Urine, Electrochemical Biosensor, Oligonucleotides, Label Free Detection, which have seven sensors such as Sarcosine Biosensor, Potentiometric Sensor, Electrochemical Biosensor, Urine Sensors, Graphene-based Electrical Sensor, Disposable Sensor Chips, and Colorimetric Biosensor in their category.

In group 8 related to Apoptosis, we have five keywords of Chemotherapy, DNA Damage, DNA Repair, Rad9, and Tumor Suppressor. Three sensors of Carbon Nanotube Biosensor, Wireless Pressure sensors, and Implantable Pressure Sensor are the important sensing technologies in this group.

### 3.4. Colorectal cancer

Finally, the colorectal cancer co-word network contains 529 nodes and 528 links. Figure 5 shows the co-occurrence network of keywords of this cancer and their connections with sensors.

**Figure 5.**
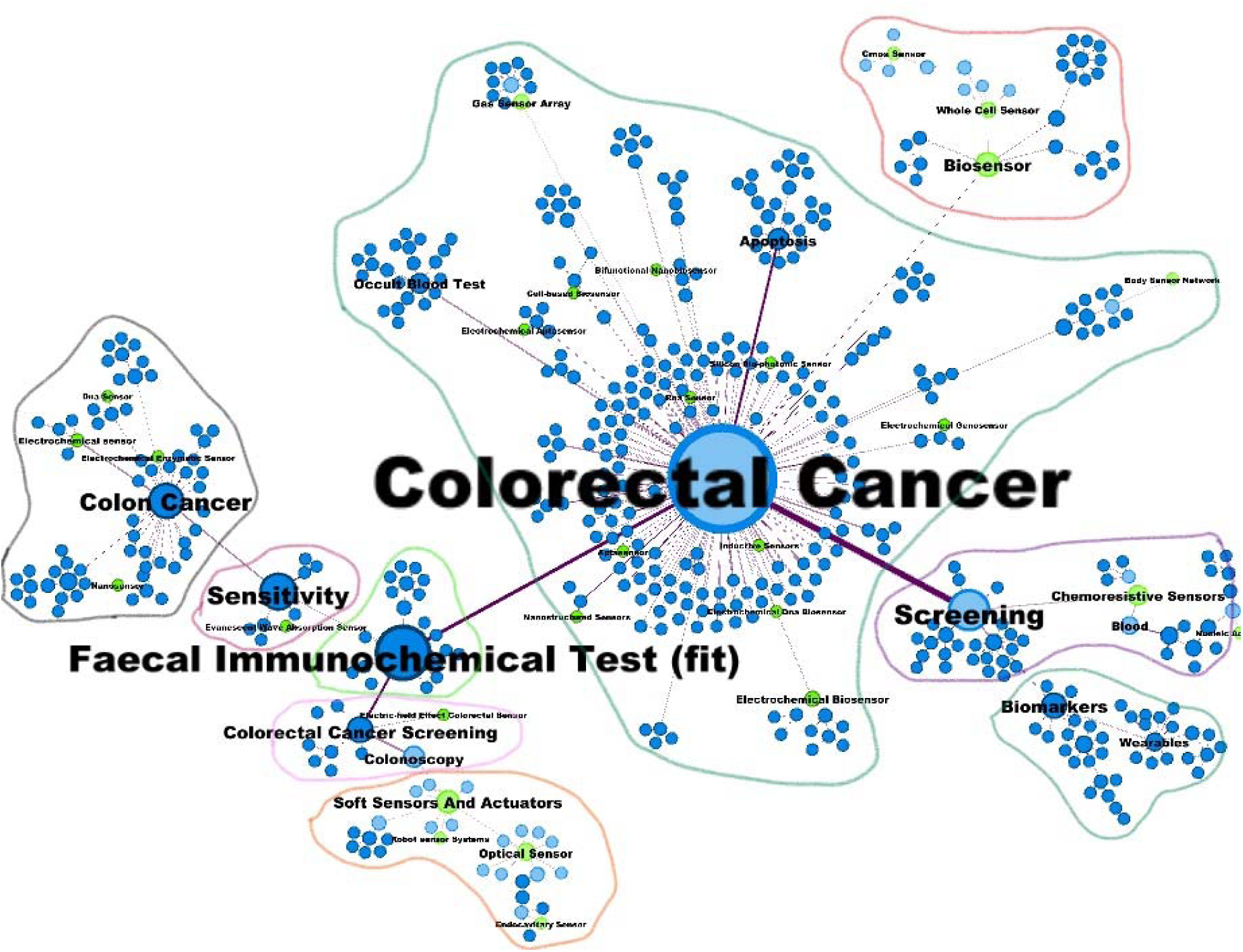
Co-word analysis map of colorectal cancer

Figure 5 shows that there are 9 groups in the colorectal cancer network. The most important nodes based on the Betweenness centrality value are colorectal cancer, Faecal Immunochemical Test, Screening, and Biosensor. Table 4A in Appendix shows the groups, core keywords of them and related sensors. Table 4A, shows the most important information about groups, core keywords and related sensors in the word co-occurrences network of the colorectal cancer. Also, there are 31 sensors interconnected to this network.

Group 1, led by Colorectal Cancer, is related to five hot topics of Apoptosis, Occult Blood Test, Precision Medicine, Social Health, and Electrochemical Biosensor. It is also illustrated that 13 sensors are interconnected to this path. Electrochemical Biosensor, Gas Sensor Array, Nanostructured Sensors, Bifunctional Nanobiosensor, and Inductive Sensors are a couple of sensors involved in creating linkage path among these nodes.

In group 2, led by Faecal Immunochemical Test (fit), we identified just 5 top keywords of Inflammatory Bowel Disease, Reg3, Quality Assurance, Advance Notification, and Letter.

Group 3 connected to Screening has two interconnected sensors including, Chemoresistive Sensors and Nucleic Acid-sensor. The most frequent keywords involved in this path group are Chemoresistive Sensors, Blood, Tumor Marker, Tissue Microarray, Faecal Occult Blood Test.

Group 4 with the head of Sensitivity is related to important keywords of Antibody, Prostatic Carcinoma, DNA, Acoustic Wave Sensor, Cancer Metastasis. Four sensors of Biosensor, Optical Sensor, Acoustic Wave Sensor, Bio-mems Force Sensor are part of this path group.

The next group, led by Colon Cancer connected to the top five keywords of Zinc, Chromogranin A, Fluorescent Sensor, making Diagnosis, and Peptide relate to four sensors of Trans-rectal Pressure Sensor Array, Chemosensor, and Fluorescent Sensor.

In Group 6, Colorectal Cancer Screening with the most important position in creating the path among different sensors and keywords is responsible for bridging Nanosensor, Flutamide Sensor, Bicalutamide Sensor and most frequent keywords of Flutamide, Nanocomposite, Voltammetry, Anticancer Drug, and Electrophilicity.

The 7^th^ group distinguished by Biomarkers includes five hot keywords of Endoscopy, Au Nps, Signal Enhancement, Erlotinib, and Tollen’s Reagent, which have two sensors: Histidine sensor and Fiber Optic Biosensor in their category.

In group 8, related to Biosensor, we have five DNA Mutation, Chemical Sensor, Urine Headspace, Colorimetric Sensor, and Tuberculosis keywords. Ten sensors of Chemical Sensor, Colorimetric Sensor, Cancer Sensor, Array-based Sensor, Temperature Sensors, Impedance-based Sensor, Attachable Gas Sensors, Hg(ii) Sensor, Eis Aptamer Sensor, and Microrna Sensor are the important sensing technologies in this group.

Group 9 led by Soft Sensors and Actuators, is related to five hot topics: Gold Nanoparticle, Immunosensor, Detection, Porcine Liver Esterase, and Fluorescent Probe. It is also illustrated that 27 sensors are interconnected to this path. Immunosensor, Near-infrared Biosensor, Resonance Sensor, Fna-based Electrochemical Sensor, Impedimetric Sensor, and Aptasensor are a couple of sensors involved in the creation of linkage path among these nodes.

According to Table 2, Biosensor is the only type of sensor that plays an essential role in all types of cancer: breast cancer, lung cancer, prostate cancer, and colorectal cancer. After that, the electrochemical sensor is in all types of cancers except lung cancer. Surprisingly, electrochemical biosensor is used in breast cancer, lung cancer, and prostate cancer research but not in colorectal cancer. Optical Sensor can also be considered one of the sensor technologies that significantly is used in three types of cancer: breast cancer, prostate cancer, and colorectal cancer. This study shows that this type of sensor is applied in more diversified approaches. Moreover, the oxygen sensor, as a type of gas sensor, is mostly applied in lung cancer and breast cancer studies due to the usage of breath analysis in the treatment process. Cmos Sensor is another technology used in two types of cancer studies, including Lung Cancer and Colorectal Cancer.

**Table 2.**
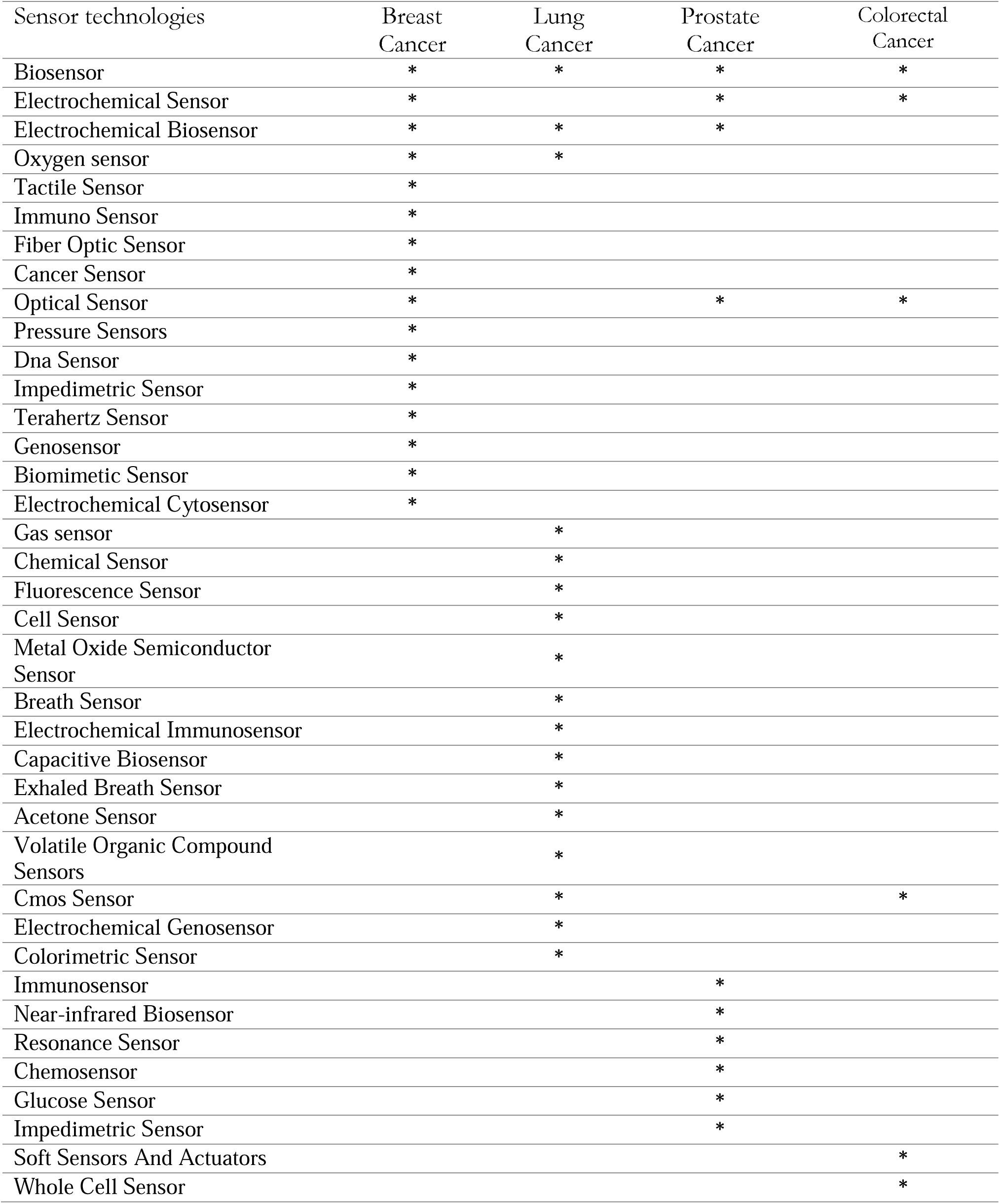
The most frequent sensors in cancer studies

| Sensor technologies | Breast Cancer | Lung Cancer | Prostate Cancer | Colorectal Cancer |
| --- | --- | --- | --- | --- |
| Biosensor | * | * | * | * |
| Electrochemical Sensor | * |  | * | * |
| Electrochemical Biosensor | * | * | * |  |
| Oxygen sensor | * | * |  |  |
| Tactile Sensor | * |  |  |  |
| Immuno Sensor | * |  |  |  |
| Fiber Optic Sensor | * |  |  |  |
| Cancer Sensor | * |  |  |  |
| Optical Sensor | * |  | * | * |
| Pressure Sensors | * |  |  |  |
| Dna Sensor | * |  |  |  |
| Impedimetric Sensor | * |  |  |  |
| Terahertz Sensor | * |  |  |  |
| Genosensor | * |  |  |  |
| Biomimetic Sensor | * |  |  |  |
| Electrochemical Cytosensor | * |  |  |  |
| Gas sensor |  | * |  |  |
| Chemical Sensor |  | * |  |  |
| Fluorescence Sensor |  | * |  |  |
| Cell Sensor |  | * |  |  |
| Metal Oxide Semiconductor Sensor |  | * |  |  |
| Breath Sensor |  | * |  |  |
| Electrochemical Immunosensor |  | * |  |  |
| Capacitive Biosensor |  | * |  |  |
| Exhaled Breath Sensor |  | * |  |  |
| Acetone Sensor |  | * |  |  |
| Volatile Organic Compound Sensors |  | * |  |  |
| Cmos Sensor |  | * |  | * |
| Electrochemical Genosensor |  | * |  |  |
| Colorimetric Sensor |  | * |  |  |
| Immunosensor |  |  | * |  |
| Near-infrared Biosensor |  |  | * |  |
| Resonance Sensor |  |  | * |  |
| Chemosensor |  |  | * |  |
| Glucose Sensor |  |  | * |  |
| Impedimetric Sensor |  |  | * |  |
| Soft Sensors And Actuators |  |  |  | * |
| Whole Cell Sensor |  |  |  | * |

## 4. Concluding remarks

The evolution of the ecosystem of sensors technology over the last few decades is unparalleled with an intensive activity of research in public and private laboratories (Andersen et al., 2004; Coccia et al., 2021; Coccia, 2005; 2015, 2017a, 2017b; Pagliaro and Coccia, 2021). Sensor technology is co-evolving with growing interactions of technological systems directed to fulfil human goals and needs and solve problems in society. Cancer is still one of the leading diseases and causes of death in the world. More than 250 types of cancers are currently known (cf., Coccia and Bellitto, 2018). Types of cancer under study here are a major cause of cancer-related deaths globally due to their difficult diagnosis in early stages resulting in late treatment. In fact, in the health domain, a major challenge is the detection of diseases using rapid and cost-effective techniques. Most of the existing cancer detection methods show poor sensitivity and selectivity and are time consuming with high cost (Mohan et al., 2022). In short, the early diagnosis is an important integral part of the process of cancer treatment. For this reason, the analysis of the role of sensor technology and research in these topics is basic in clinical diagnosis and early treatment for patients for reducing the mortality worldwide.

The task of this study was to do an exploratory analysis on the role of sensor technology in cancer research to see possible new directions for improving diagnosis and therapeutic treatments. The results of this analysis are:

1. Biosensor is the only type of sensor that plays an essential role in all types of four cancer under study.
2. Electrochemical sensor is in all types of cancer except lung cancer.
3. Electrochemical Biosensor is used in breast cancer, lung cancer, and prostate cancer research but not colorectal cancer.
4. Optical Sensor is a technology used in three types of cancer: breast cancer, prostate cancer, and colorectal cancer.
5. Oxygen sensor has a role in lung cancer and breast cancer studies due to the usage for breath analysis in the treatment process.
6. Finally, Cmos Sensor is another technology used in two types of cancer studies, including Lung Cancer and Colorectal Cancer.

Overall, then, results suggest that new directions, such as optical biosensors that are rapid, real-time, and portable technology, have a low detection limit and a high sensitivity, and have a great potential for diagnosing various types of cancer. Optical biosensors can detect cancer in a few million malignant cells, in comparison to conventional diagnosis techniques that use 1 billion cells in tumor tissue with a diameter of 7 nm–10 nm (traditional methods that are also costly, inconvenient, complex, time consuming, and require technical specialists; Kaur et al., 2022). Moreover, the cancer biomarkers using luminescence and electrochemical Metal-organic framework sensors have been opening the way for personalized patient treatments and the development of new cancer-detecting devices (Mohan et al., 2022). The challenge of sensor technology in cancer research is the developing of simple, reliable and sensitive point-of-care testing biosensor for cancerous exosomes detection to early cancer diagnosis and prognosis. The biosensor could also avoid the influence of the external environment, including surrounding light and temperature(Zou et al., 2022).

These conclusions are, of course, tentative. Although this study has provided some interesting, albeit preliminary results, it has several limitations. First, a limitation of this study is that sources understudy may only capture certain aspects of the ongoing dynamics of sensor research and technology in cancer research. Second, there are multiple confounding factors that could have an important role in the interaction between sensor technology and cancer research for diagnosis to be further investigated, such as high R& D investments, collaboration intensity, openness, intellectual property rights, etc. (Roshani et al., 2021). Third, the computational and statistical analyses in this study focus on data in a specific period and should be extended to other periods. Forth, sensor research associated with cancer studies change their borders during the evolution of science, such that the identification of stable technological trajectories and new patterns in the evolution of sensors in cancer research is a non-trivial exercise.

To conclude, future research should consider new data when available, and when possible, apply new approaches to reinforce proposed results. Despite these limitations, the results presented here clearly illustrate the evolutionary paths of main sensor technologies that have a great potential as a powerful tool in future diagnosis of cancer but we also need a detailed examination of other aspects and factors for supporting appropriate strategies of research and innovation policy, and management of technology to foster the technology transfer of sensor in cancer research for improving diagnosis and at the same time reducing, as far as possible, world-wide mortality of cancer ins society.

## Data Availability

All data produced in the present study are available upon reasonable request to the authors

## Appendix

**Table 1A.**
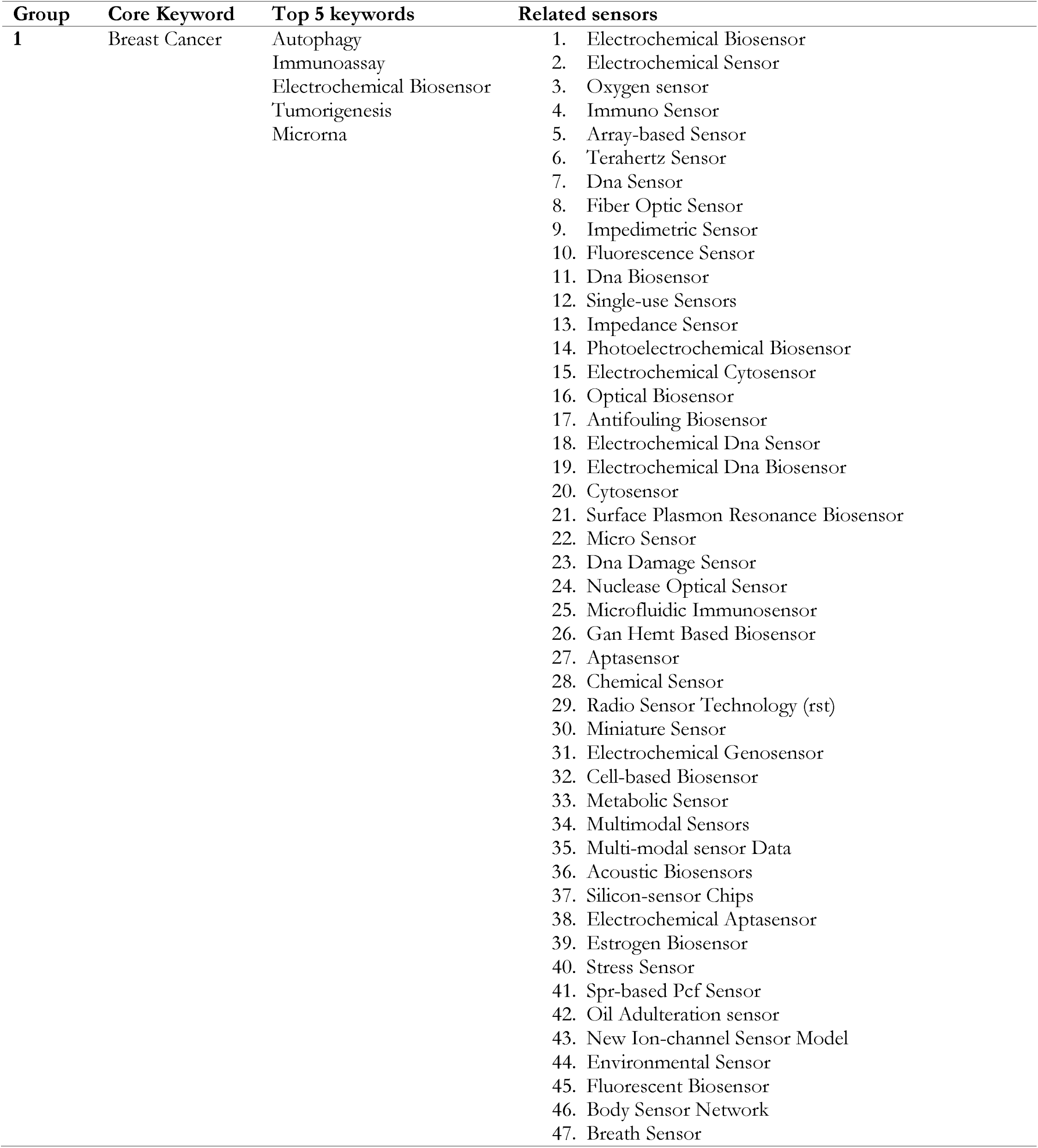
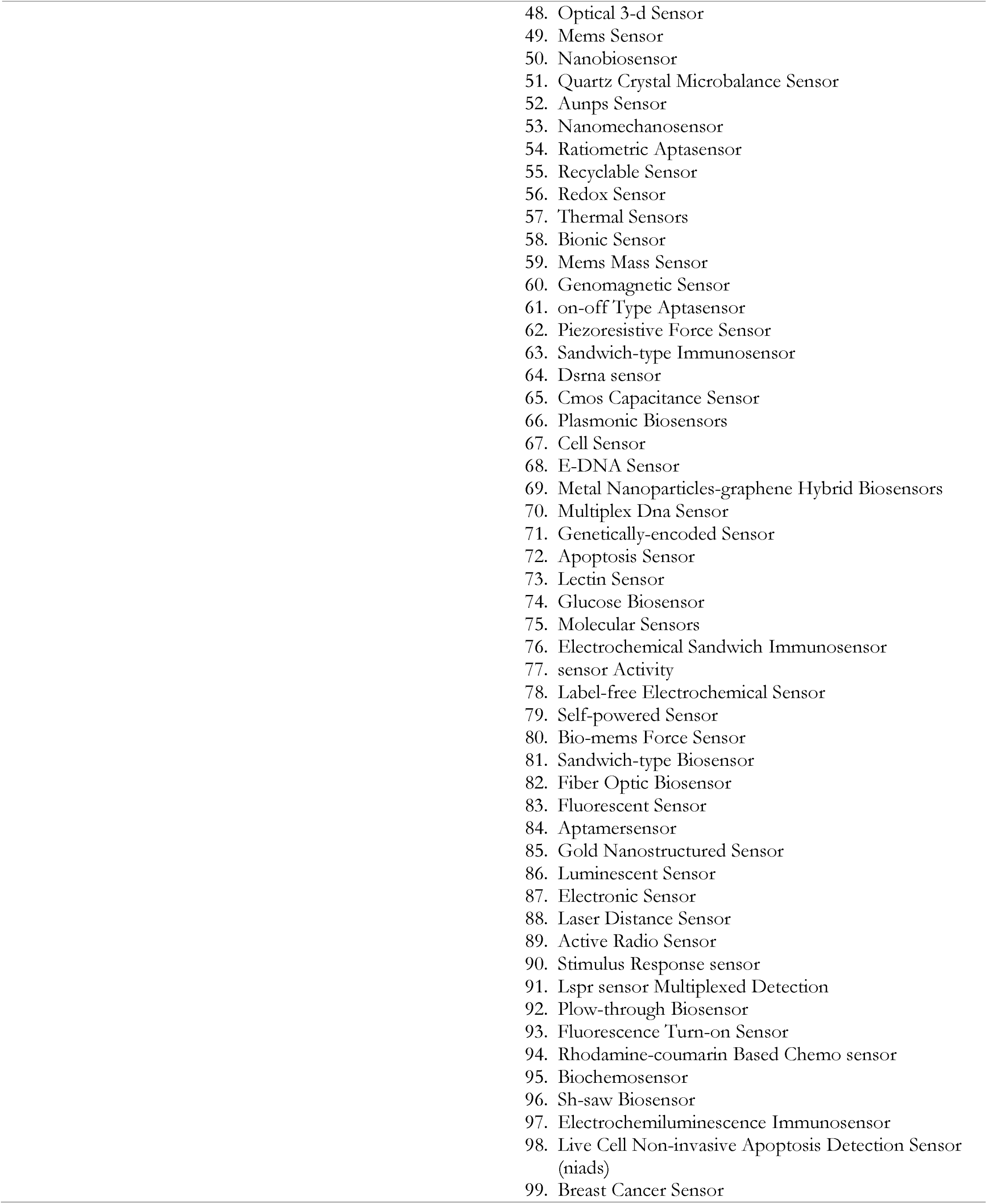
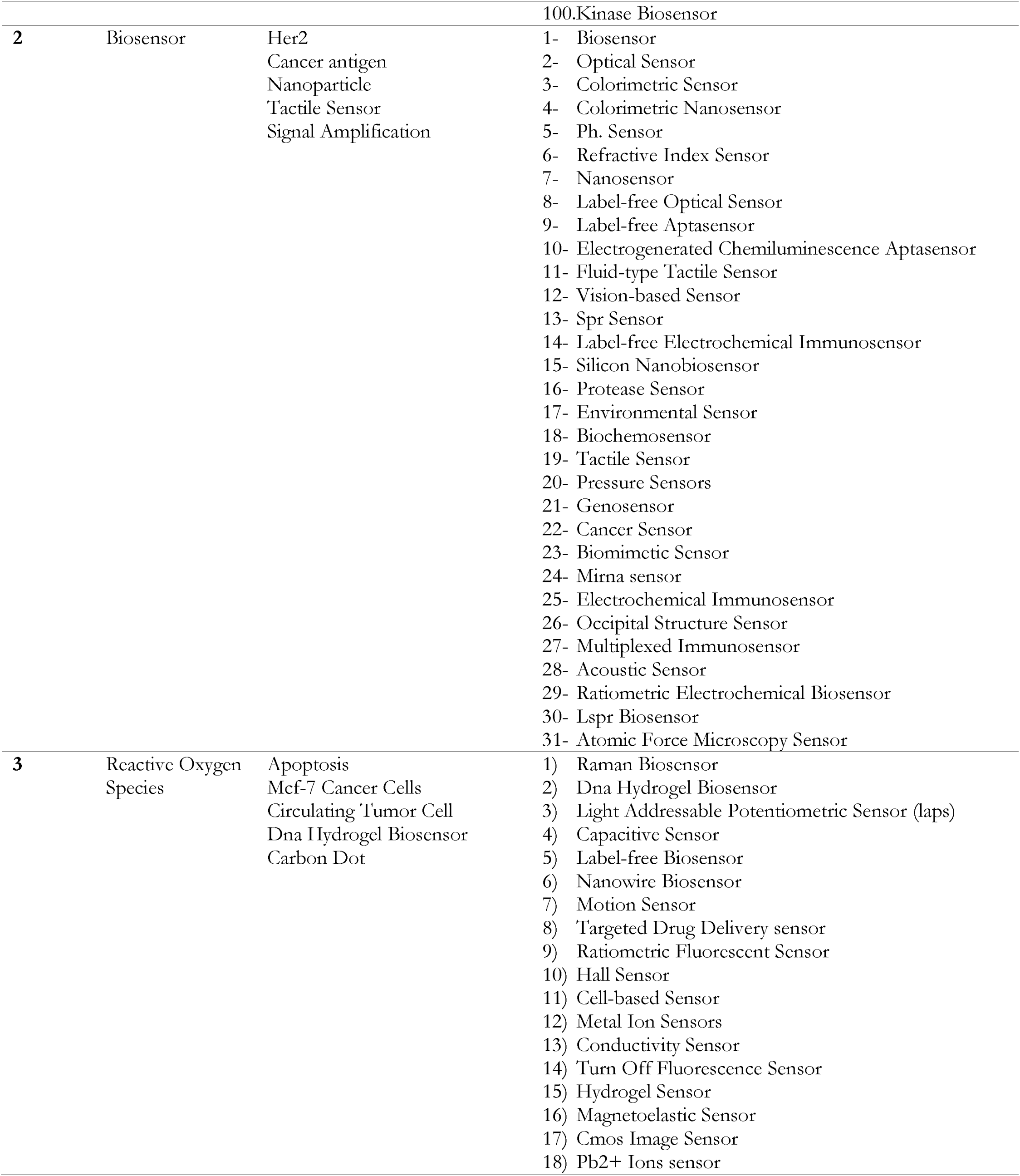
Groups, Core keywords and related sensors in the breast cancer network

**Table 2A.**
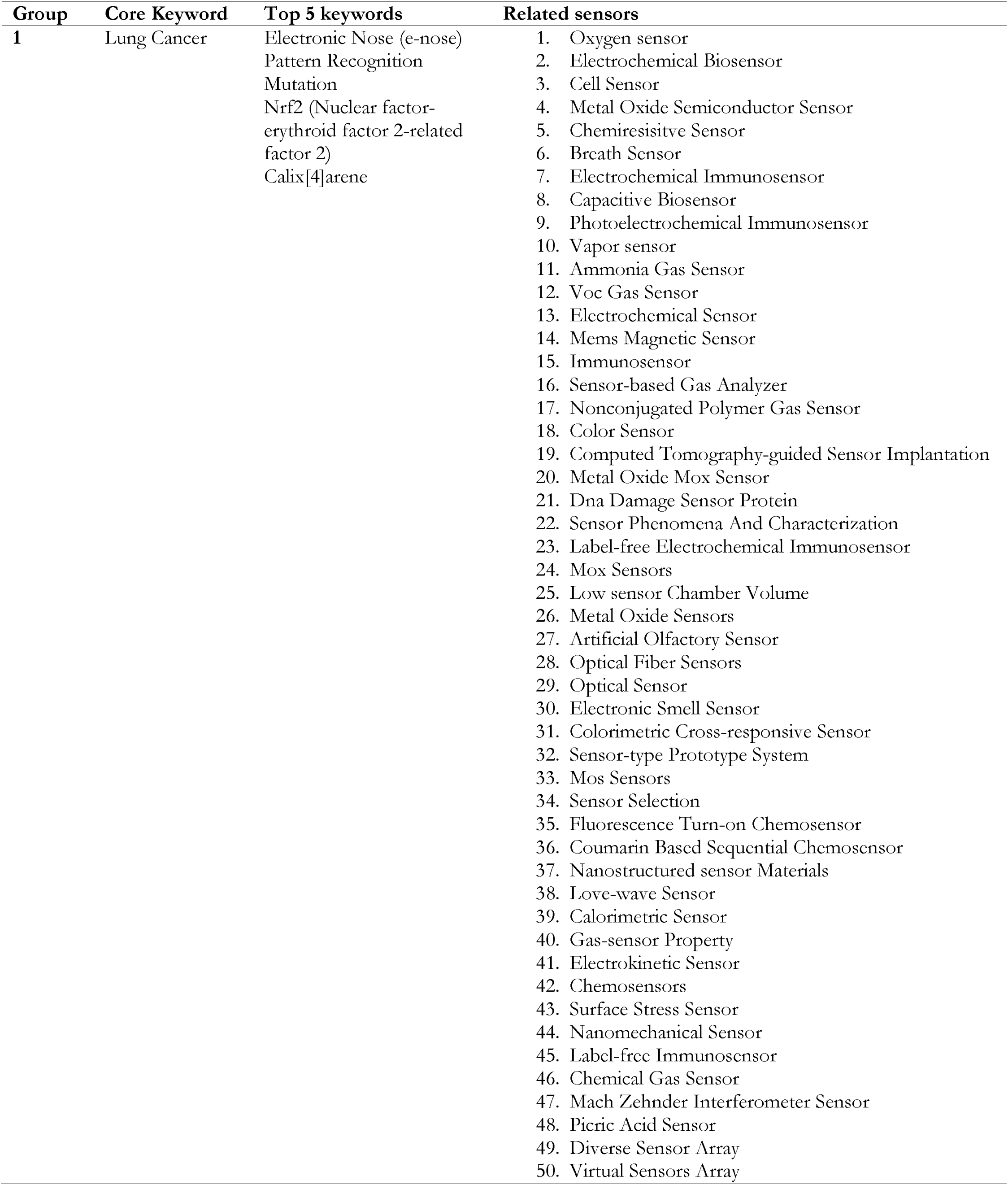
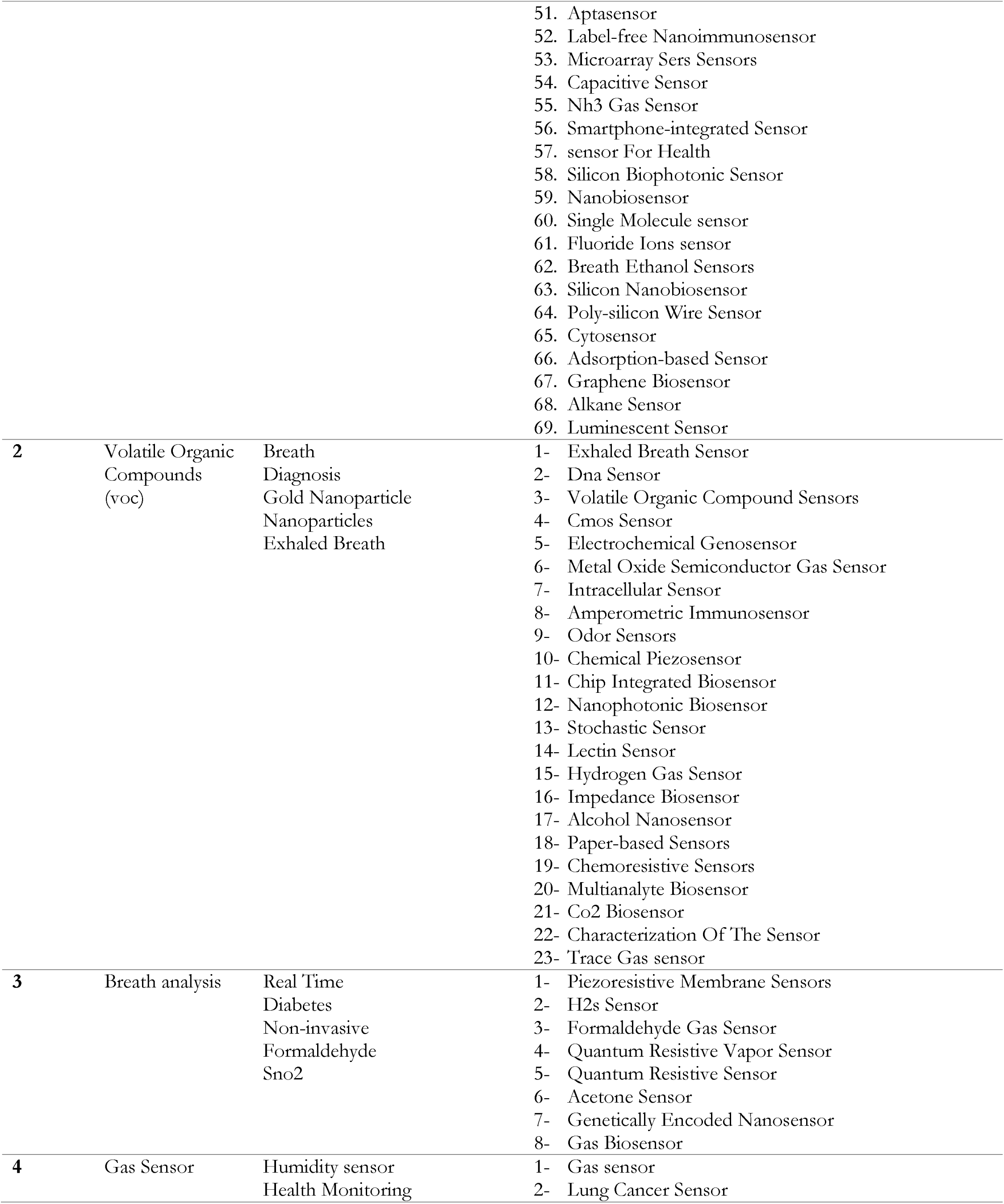
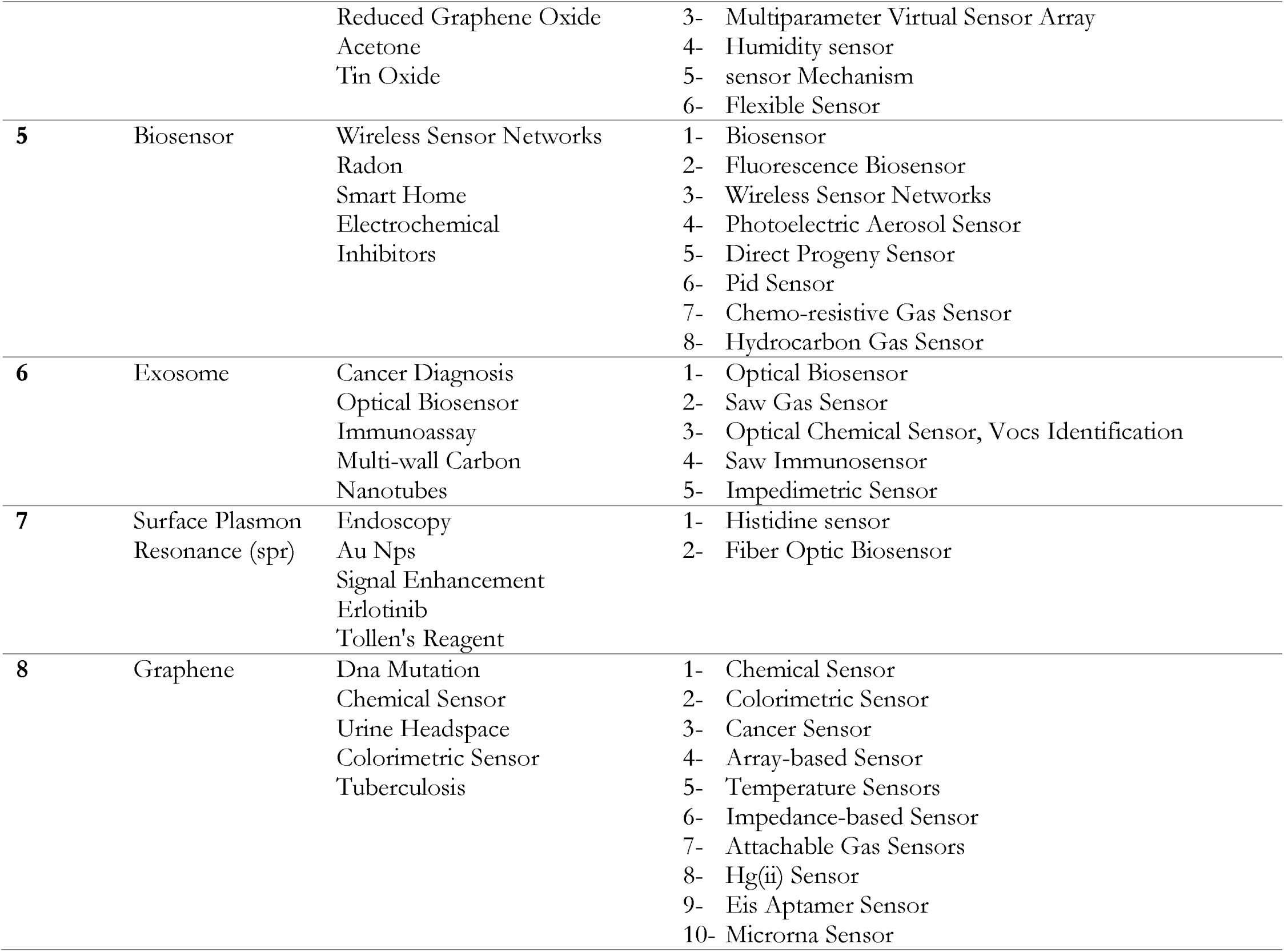
Groups, Core keywords and related sensors in the lung cancer network

**Table 3A.** Groups, Core keywords and related sensors in the Prostate cancer network

| Group | Core Keyword | Top 5 keywords | Related sensors |
| --- | --- | --- | --- |
| 1 | Prostate cancer | Gold Nanoparticle<br>Immunosensor<br>Detection<br>Porcine Liver Esterase<br>Fluorescent Probe | 5. Immunosensor<br>6. Near-infrared Biosensor<br>7. Resonance Sensor<br>8. Fna-based Electrochemical Sensor<br>9. Impedimetric Sensor<br>10. Aptasensor<br>11. Piezoelectric Sensor<br>12. Boronate Sensor<br>13. Zinc sensor<br>14. Nanopore-based Sensor<br>15. Nanowire Biosensor<br>16. Microsensors<br>17. Voltammetric Sensor<br>18. Hall Sensor<br>19. Electrochemical Aptasensor<br>20. Tactile Sensor<br>21. Array-based Sensor<br>22. Non-enzymatic Sensor<br>23. Electrochemical Aptamer Sensors<br>24. Ratiometric Aptasensor<br>25. Light Addressable Potentiometric Sensor (laps)<br>26. Camera Sensor<br>27. Fluorescence Gas-sensory Arrays |
| 2 | Activated Protein Kinase (Ampk) | Apoptosis<br>Lkb1<br>Lung Cancer<br>Mtorc1<br>Castrate-resistant | 1. Label-free Nanoimmunosensor |
| 3 | Prostate Specific Antigen (psa) | Aptamer<br>Prostate Cancer Diagnosis<br>Cancer Biomarker<br>Silicon Nanowire | 1. Glucose Sensor<br>2. Ph Sensor<br>3. Colorimetric Sensor<br>4. Quantum Resistive Sensor<br>5. Label-free Biosensors<br>6. Love Wave Sensor<br>7. Biomarker sensor<br>8. Photonic Sensor<br>9. Biomarker Pressure Sensor<br>10. Carbon Nanotube Sensors<br>11. Ion Selective Sensors<br>12. Rna Biosensor<br>13. Affinity Sensor<br>14. Electrical Conductivity Sensor<br>15. Conductivity Sensor<br>16. Memristive Biosensors<br>17. Ring Resonator Biosensor<br>18. Nanobiosensor<br>19. Metal Nanoparticles-graphene Hybrid Biosensors<br>20. Impedimetric Immunosensor<br>21. Single-molecule Sensor |
|  |  |  | 22. Sandwich-type Electrochemical Immunosensor |
| 4 | Biosensor | Antibody<br>Prostatic Carcinoma<br>Dna<br>Acoustic Wave Sensor<br>Cancer Metastasis | 1. Biosensor<br>2. Optical Sensor<br>3. Acoustic Wave Sensor<br>4. Bio-mems Force Sensor |
| 5 | Prostate | Zinc<br>Chromogranin A<br>Fluorescent Sensor<br>Imaging Diagnosis<br>Peptide | 1. Trans-rectal Sensor<br>2. Pressure Sensor Array<br>3. Chemosensor<br>4. Fluorescent Sensor |
| 6 | Electrochemical Sensor | Flutamide<br>Nanocomposite<br>Voltammetry<br>Anticancer Drug<br>Electrophilicity | 1. Electrochemical Sensor<br>2. Nanosensor<br>3. Flutamide Sensor<br>4. Bicalutamide Sensor |
| 7 | Sarcosine | Urine<br>Electrochemical Biosensor<br>Oligonucleotides<br>Label Free Detection | 1. Sarcosine Biosensor<br>2. Potentiometric Sensor<br>3. Electrochemical Biosensor<br>4. Urine Sensors<br>5. Graphene-based Electrical Sensor<br>6. Disposable Sensor Chips<br>7. Colorimetric Biosensor |
| 8 | Apoptosis | Chemotherapy<br>Dna Damage<br>Dna Repair<br>Rad9<br>Tumor Suppressor | 1. Carbon Nanotube Biosensor<br>2. Wireless Pressure Sensor<br>3. Implantable Pressure Sensor |

**Table 4A.**
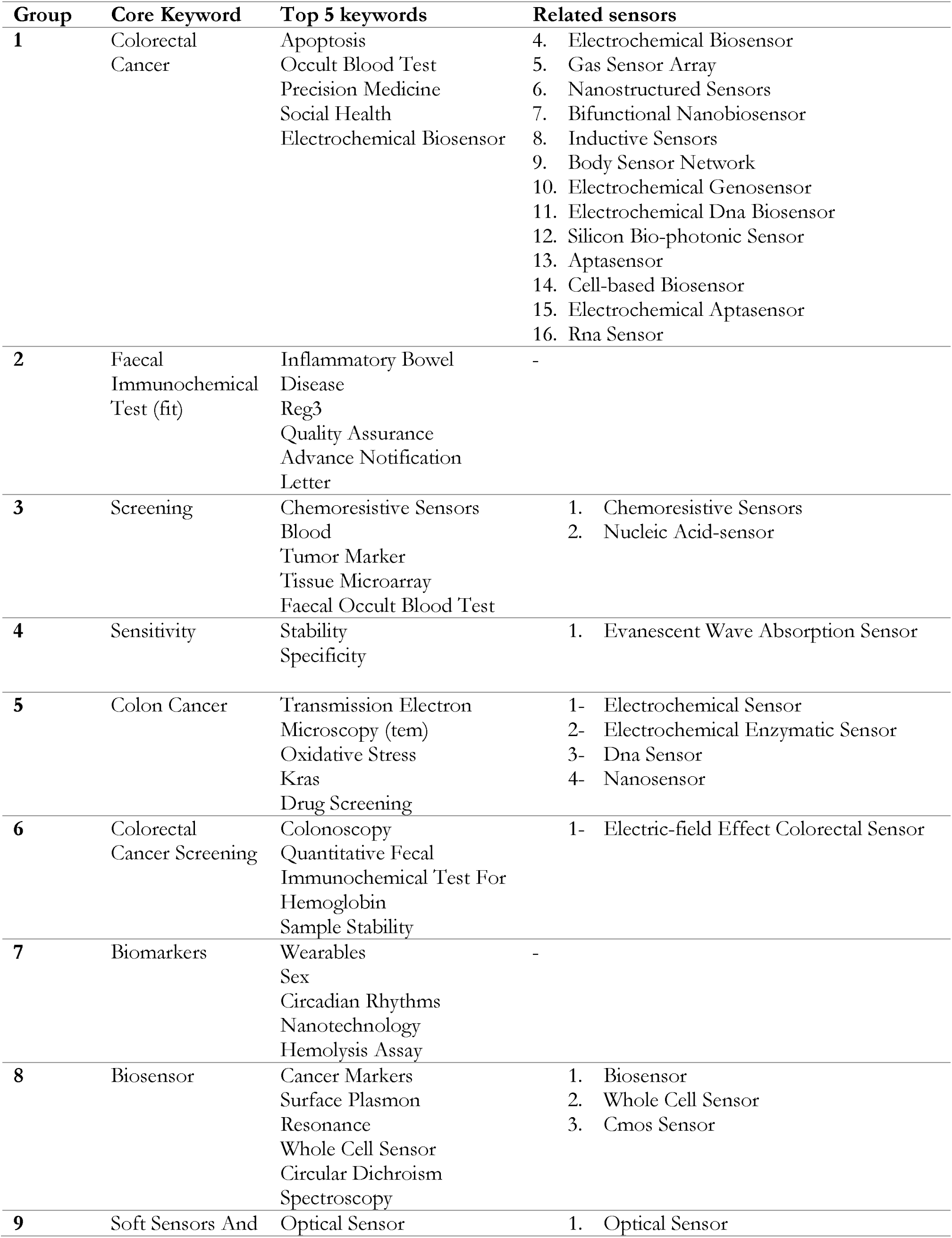
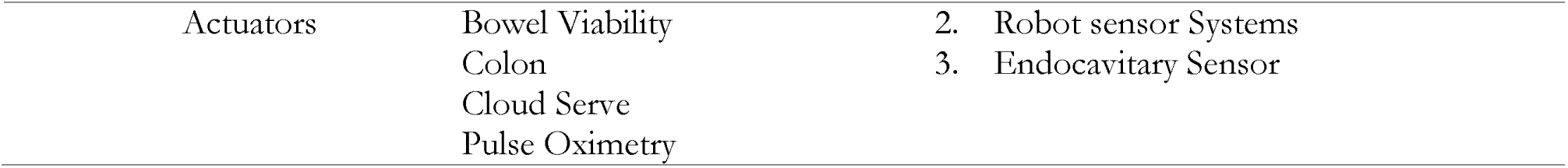
Groups, Core keywords and related sensors in the colorectal cancer network

